# Estimating dengue transmission intensity from serological data: a comparative analysis using mixture and catalytic models

**DOI:** 10.1101/2021.10.29.21259708

**Authors:** Victoria Cox, Megan O’Driscoll, Natsuko Imai, Ari Prayitno, Sri Rezeki Hadinegoro, Anne-Frieda Taurel, Laurent Coudeville, Ilaria Dorigatti

## Abstract

**Background:** Dengue virus (DENV) infection is a global health concern of increasing magnitude. To target intervention strategies, accurate estimates of the force of infection (FOI) are necessary. Catalytic models have been widely used to estimate DENV FOI and rely on a binary classification of serostatus as seropositive or seronegative, according to pre-defined antibody thresholds. Previous work has demonstrated the use of thresholds can cause serostatus misclassification and biased estimates. In contrast, mixture models do not rely on thresholds and use the full distribution of antibody titres. To date, there has been limited application of mixture models to estimate DENV FOI.

**Methods:** We compare the application of mixture models and time-constant and time-varying catalytic models to simulated data and to serological data collected in Vietnam from 2004 to 2009 (N ≥ 2178) and Indonesia in 2014 (N = 3194).

**Results:** The simulation study showed greater estimate bias from the time-constant and time-varying catalytic models (FOI bias = 1.3% (0.05%, 4.6%) and 2.3% (0.06%, 7.8%), seroprevalence bias = 3.1% (0.25%, 9.4%) and 2.9% (0.26%, 8.7%), respectively) than from the mixture model (FOI bias = 0.41% (95% CI 0.02%, 2.7%), seroprevalence bias = 0.11% (0.01%, 3.6%)). When applied to real data from Vietnam, the mixture model frequently produced higher FOI and seroprevalence estimates than the catalytic models.

**Conclusions:** Our results suggest mixture models represent valid, potentially less biased, alternatives to catalytic models, which could be particularly useful when estimating FOI and seroprevalence in low transmission settings, where serostatus misclassification tends to be higher.

**Author summary:** Characterising the transmission intensity of dengue virus in different geographic areas over time is essential to understand who is at greatest risk of infection, and to inform the implementation of interventions, such as vector control and vaccination. It is therefore important to understand how methodological differences and model choice may influence estimates of transmission intensity. We compared the application of catalytic and mixture models to calculate the force of infection (FOI) of dengue virus from antibody titre data. We observed greater bias in FOI estimates obtained from catalytic models than from mixture models in areas where the transmission intensity was low. In high transmission intensity areas, catalytic and mixture models produced consistent estimates. Our results indicate that in low transmission settings, when antibody titre data are available, mixture models could be preferential to estimate dengue virus FOI.

## Introduction

Dengue fever is caused by infection with one or more of four antigenically distinct serotypes of dengue virus (DENV1-4); a *Flavivirus* carried by *Aedes* mosquitoes (1,2). DENV infects approximately 105 million people each year (3), primarily in tropical and sub-tropical regions. The geographical range of DENV is increasing (1,4,5) and it is expected that the spread of dengue will be influenced by rising global temperatures and increasing urbanisation (1,6). Intervention measures to date rely essentially on vector control due to the absence of antiviral treatment, challenges in the use of the first licensed dengue vaccine for widespread dengue prevention and control (7), as well as in the use of rapid diagnostic tests for screening (8). The current and expected future burden of dengue on health-systems is therefore high, demonstrating a continuing need for increased understanding of DENV transmission.

Estimating epidemiological parameters such as the force of infection (FOI, the per capita rate at which a susceptible person is infected) and population seroprevalence (the proportion of people in a population exposed to a virus, as determined by the detection of antibodies in the blood) allow us to gain insights into the subsets of populations most at risk of infection and disease (9), to assess the predicted impact of an intervention strategy (10), and to inform public health policy (11,12).

Both the FOI and seroprevalence can be estimated using mathematical models calibrated to serological data measuring IgG antibody levels (also called titres) from blood samples. IgG titres are obtained using Enzyme-Linked Immunosorbent Assays (ELISAs) and are often classified into qualitative, binary test results (seropositive or seronegative) based on the manufacturer’s threshold.

Catalytic models, first proposed in 1934, estimate disease FOI from age-stratified serological or case notification data (13). In these models, large rates of increase in seroprevalence between individuals who are age *a* versus age *a+1* are explained by high age-specific FOI (assuming the FOI is constant in time) or high time-specific FOI experienced by individuals of all ages during the period *a* to *a+1* years ago (14). Catalytic models have been used extensively for measles (15), rubella (16), Hepatitis A (17), Chagas disease (18), and DENV (12,14,19–21). Whilst commonly used, Bollaerts *et al*. and Vink *et al*. (22,23) suggest that catalytic models risk generating biased estimates due to data-loss and/or mis-classification. For example, samples with titres greater than the seronegative threshold but lower than the seropositive threshold are classified as ‘equivocal’ and discarded from the analysis. Furthermore, titre levels of seropositive individuals in a given population may be affected by factors including host response, the degree of exposure to the pathogen and infection timing, which could lead to misclassification.

Mixture models are flexible statistical models that can be applied to continuous data from different clusters or populations, called components. Mixture models can therefore be applied to the absolute antibody titre values in serology datasets, rather than to the counts of titres in each of two classes (seropositive/seronegative) as is necessary for catalytic models (22). The components’ distributions and their defining parameters (e.g., the mean titre of each component distribution) are inferred from a fitted mixture model which is used to estimate the FOI and population seroprevalence (22,24). To date, mixture models have been applied to serological data to estimate the seroprevalence of infectious diseases such as parvovirus B19 and rubella in England (25,26), human papillomavirus in the Netherlands (23), measles in Italy (27), and a selection of arboviruses inlcuding DENV in Zambia (28). In addition, mixture models have been used to develop a framework capable of distinguishing between primary and post-primary DENV infections (29) and to estimate the FOI of Varicella-Zoster virus in Belgium (22). However, these methods have not been used to estimate DENV FOI. Here, we present the results of a simulation study and a comparison of the DENV trasmission intensity estimates obtained from mixture and catalytic models applied to age-stratified serological datasets from Vietnam (N ≥ 2178, for years 2004-2009) and Indonesia (N = 3194, for 2014).

## Methods

### Data

#### Age-stratified seroprevalence data

DENV IgG data were collected in Long Xuyen, Vietnam, during a prospective epidemiological study that was conducted to assess the suitability of the area for future CYD-TDV vaccine efficacy trials, as described previously (30). Samples were collected from children under 11 years old in 2004 and then from children under 15 years old during September to February in 2004-2005, 2005-2006, 2006-2007, 2007-2008 and 2008-2009 (Datasets A-1 to A-6, Table 1). The titres were measured using in-house ELISA assays (Arbovirus Laboratory of Pasteur Institute, Ho Chi Minh City).

**Table 1:** Description of the datasets used in the analyses. Summary statistics including notation, region, the assay used, the year of testing, the age range of the children participating to the study and the sample sizes.

| Dataset | Region | Assay type | Year | Age<br>Range | Sample Size |
| --- | --- | --- | --- | --- | --- |
| A-1 | Long Xuyen, Vietnam | IgG ELISA | 2004 | 3-10 | 2,178 |
| A-2 | Long Xuyen, Vietnam | IgG ELISA | 2004-2005 | 3-13 | 3,681 |
| A-3 | Long Xuyen, Vietnam | IgG ELISA | 2005-2006 | 3-14 | 3,727 |
| A-4 | Long Xuyen, Vietnam | IgG ELISA | 2006-2007 | 3-14 | 3,651 |
| A-5 | Long Xuyen, Vietnam | IgG ELISA | 2007-2008 | 4-14 | 2,959 |
| A-6 | Long Xuyen, Vietnam | IgG ELISA | 2008-2009 | 5-14 | 2,249 |
| B | 30 urban subdistricts,<br>Indonesia | IgG ELISA | 2014 | 1-18 | 3,194 |
| C | Simulated data | Simulated | N/A | 1-18 | 3,194 |

DENV IgG data from 30 urban subdistricts in Indonesia were collected from children under 18 years old as part of a cross-sectional seroprevalence survey in 2014 (31) (Dataset B). The total number of participants was 3,194 (see Supplementary Data for subdistrict information). IgG titres were measured using the commercial Panbio® Dengue IgG Indirect ELISA kit.

#### Simulated datasets

We simulated 200 antibody titre datasets (Dataset C), with the same age-distribution and sample size as the Indonesian seroprevalence survey data (Dataset B). For each simulation the distributions used for sampling seronegative and seropositive antibody titres were randomly selected from a normal, gamma or Weibull distribution. This gave 9 possible combinations of distribution pairs for seronegative and seropositive antibody titre values per simulation. Normal, gamma and Weibull distributions were chosen based on preliminary work where mixture models applied to our antibody titre datasets most commonly fitted one of these three distributions. Parameter values were randomly drawn from uniform distributions with limits as shown in S2 Table. The serostatus of each individual was drawn from a Bernoulli distribution with probability 1-*e*^-*λ a*^, where *a* is the age of the individual and *λ* is the FOI. FOI was assumed to be constant with age and time. Titre values for each individual were subsequently randomly drawn from the respective component distributions. The analysis was conducted in the statistical programming language R (32).

#### Ethics statement

Ethical approval for the secondary analysis of the age-stratified seroprevalence datasets was granted by the Imperial College Research Ethics Committee (Approval Reference 21IC7066).

#### Catalytic model

##### Data preparation

Catalytic models rely on data that are binarily classified as seropositive or seronegative. For Datasets A-1 to A-6, a background/control titre (t) was measured for each assay. An individual titre was classified as seronegative if ≤t and seropositive otherwise. For Dataset B, samples with titres ≤ 9 PanBio units were classified as seronegative and ≥11 as seropositive. Titres >9 and <11 were discarded (28 out of 3,194 samples). For simulated Dataset C, titres were classified as seronegative if they were ≤X and seropositive if they were ≥Y. X and Y are thresholds that were optimised using the ‘true’ simulated serostatuses: the *optim* function in R, using the Nelder Mead algorithm, was used to calulate the X and Y values per simulated dataset resulting in the fewest titre misclassifications. The mean number of samples discarded (titre value >X and <Y) per simulation was 1.06 out of 3194 (median = 0, interquartile range (IQR) = 1, range = 0 to 19). The mean percentage of titres misclassified across the simulations was 2.4% (median = 0.6%, IQR = 2.0%, range = 0% to 69.4%).

##### Parameter estimation

We used the same catalytic model developed in Rodriguez-Barraquer *et al*., 2011 (33). The proportion of seropositive individuals in age group *i (π*(*a*_*i*_)), was estimated as in Equation 1, where, *λ*(*a*_*i*_) denotes the FOI, i.e. the per-capita rate of infection experienced by individuals in age group *i*, and a denotes the age (14).

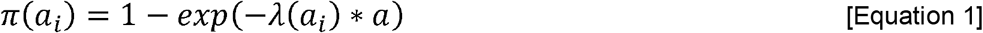

A binomial log-likelihood was assumed for the FOI (Equation 2), where *N*_*i*_ is the total number of individuals and *X*_*i*_ is the number of seropositive individuals in age group *i* (19). The maximum likelihood estimate of the FOI parameter is estimated as shown in Equation 2 using the *optim* function in R.

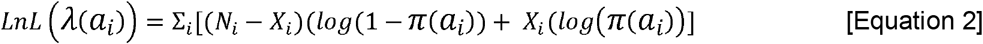

When we assume the FOI is time-varying, the estimated *λ*(*a*_*i*_) values were averaged across the age groups (number of age groups = *n*) to give the mean yearly FOI experienced over the study period (Equation 3). When we assume the FOI is constant with time, *λ*(*a*_*i*_) equals, *λ*.

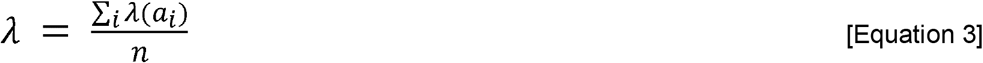

We estimated 95% Confidence Intervals (CI) using a bootstrap method, where the data were sampled with replacement 1000 times. The 95% CI was given by the 2.5% to 97.5% quantiles of the estimates from the bootstrap samples.

## Mixture model

### Applying the mixture models to the titre distributions

Mixture models were applied to the bimodal distribution of individual antibody titres as described in Bollaerts *et al*., 2012 and Hens *et al*., 2012. The model defines the distribution of the log(titres + 1) (*z*) as a mixture of two distinct distributions: one for susceptible individuals (seronegative, *z*_*s*_) and one for individuals who have been previously infected (seropositive, *z*_*I*_). The two-component mixture model is represented by:

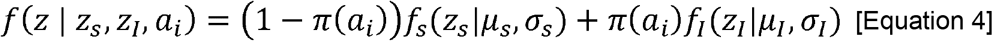

where *f*_*s*_ and *f*_*I*_ represent the probability density function of the seronegative and seropositive components, respectively, and where *μ* and *σ* represent the mean and standard deviation of each component. The age-dependent mean log(titre+1) (*μ*(*a*_*i*_)) is calculated as in Equation 5, which corresponds to an expression for the age-specific seroprevalence (*π*(*a*_*i*_)) (Equation 6).

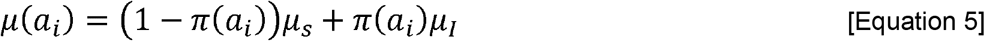

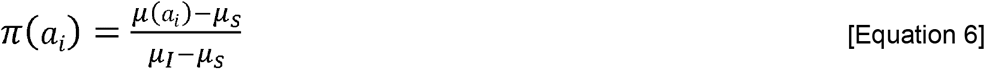

The age-dependent FOI, *λ*(*a*_*i*_), can be derived from the seroprevalence as described in Equation 7 (22) and in turn can be expressed as a function of the underlying antibody titre distribution as shown in Equation 8, where *μ*’(*a*_*i*_) represents the derivative of the age-specific mean log(titre+1). The age-dependent FOI can be averaged across the age groups to give the total FOI, *λ* (Equation 3).

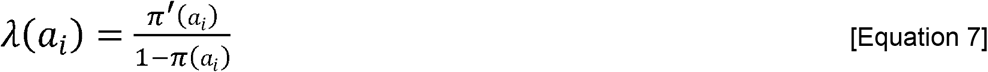

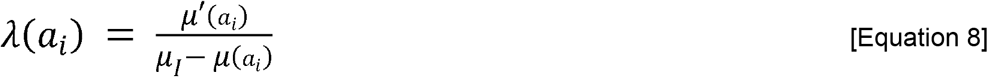

The *mixdist* R package (34) was used to fit the mixture models to the titre data using an Expectation Maximisation (EM) algorithm. The package was adapted to allow fitting of different distributions for the seronegative and seropositive titre components: normal, gamma and Weibull distributions were fitted for each component, giving 9 possible combinations. The model giving the best fit to the titre distribution (determined by minimising the log-likelihood ratio statistic) among the 9 combinations tested was chosen and its estimates of the means (*μ*_*s*_ and *μ*_*I*_) and standard deviations (*σ*_*S*_ and *σ*_*I*_) for the seronegative and seropositive components were extracted (and for Dataset C they were compared, as well as the FOI, λ, against the true simulated parameter values). We explored multiple parameterisations, including fixing the standard deviation of the two mixture components. For the Vietnamese datasets, we optimised the model having constrained the standard deviation of the seropositive component to multiple different values (for Dataset A-4 we set *σ*_*I*_ equal to all values from 0.02 to 0.08 in steps of 0.01, for the other five Datasets we set *σ*_*I*_ equal to all values from 0.05, to 0.15 in steps of 0.01). For the Indonesian dataset (Dataset B) the standard deviations of both components were constrained (*σ*_*S*_ was set equal to 0.10 to 0.15 in steps of 0.001, and *σ*_*I*_ was set equal to 0.15 to 0.3 in steps of 0.05).

### Parameter estimation

The distribution mean, *μ*(*a*), was estimated by least-squares regression using a monotonically increasing P spline fitted to the age-stratified log(titre+1) data (22,24,35). Equally spaced cubic polynomial segments (degree = 3) made up the spline. The optimal smoothing parameter (*α*) and number of segments (knots) were determined using the Bayesian Information Criterion, having explored combinations of *α* values (set equal to 0.001, 0.01, 0.1, 0.5, 1, 5, 10, 50, 100, 500) and knots (set equal to values in the sequence: 5 to the maximum number of x-axis age categories, step size = 1). The spline was fitted using the *mpspline*.*fit* function from the *serostat* R package (36). The *μ’*(*a*) term for the FOI estimation was calculated by taking the gradient of the fitted spline at each age. The 95% CI around the parameter estimates were calculated using a boostrap method, as described above. Code to recreate the simulated datasets and to run the mixture model analysis are available at: https://github.com/Tori-Cox/Mixture-catalytic-models.

## Results

### Simulated data

The mixture model identified the correct distributions used to simulate both seropositive and seronegative titres in 50% (100/200) of simulations, one of the two distributions in 42.5% (85/200) of simulations and did not correctly identify either distribution in 7.5% (15/200) of the simulations. In all 15 simulations where neither distribution was correctly identified, the seronegative titre values were drawn from the Weibull distribution (S3 Table). The estimated 95% CIs contained the true parameter values used to simulate the data in 87% (174/200), 92% (184/200), 80.5% (161/200) and 90.5% (181/200) of simulations for *μ*_*s*_, *μ*_*I*_, *σ*_*S*_ and *σ*_*I*_, respectively (S5 Figure).

The mixture model correctly estimated the FOI for 99% (198/200) and the seroprevalence for 97.5% (195/200) of simulations. The time-varying catalytic model correctly estimated the FOI for 98% (196/200) and the seroprevalence for 85% (170/200) of simulations. The time-constant catalytic model correctly estimated the FOI and seroprevalence for 8.5% (17/200) and 16.5% (33/200) of simulations, respectively. The estimates were categorised as correct if the estimated 95% CIs contained the true values. It should be noted that the time-varying catalytic model produced wider CIs compared to when assuming a constant-in-time FOI (Figure 1). Bias was calculated as the estimated parameter values minus the true parameter values. Absolute bias in the FOI estimates (0.41% (95% CI 0.02%, 2.7%), 1.3% (0.05%, 4.6%) and 2.3% (0.06%, 7.8%) for the mixture, time-constant and time-varying catalytic models, respectively) and the seropreavelance estimates (0.11% (0.01%, 3.6%), 3.1% (0.25%, 9.4%) and 2.9% (0.26%, 8.7%)) was smaller for the mixture model estimates (Figure 1).

**Figure 1:**
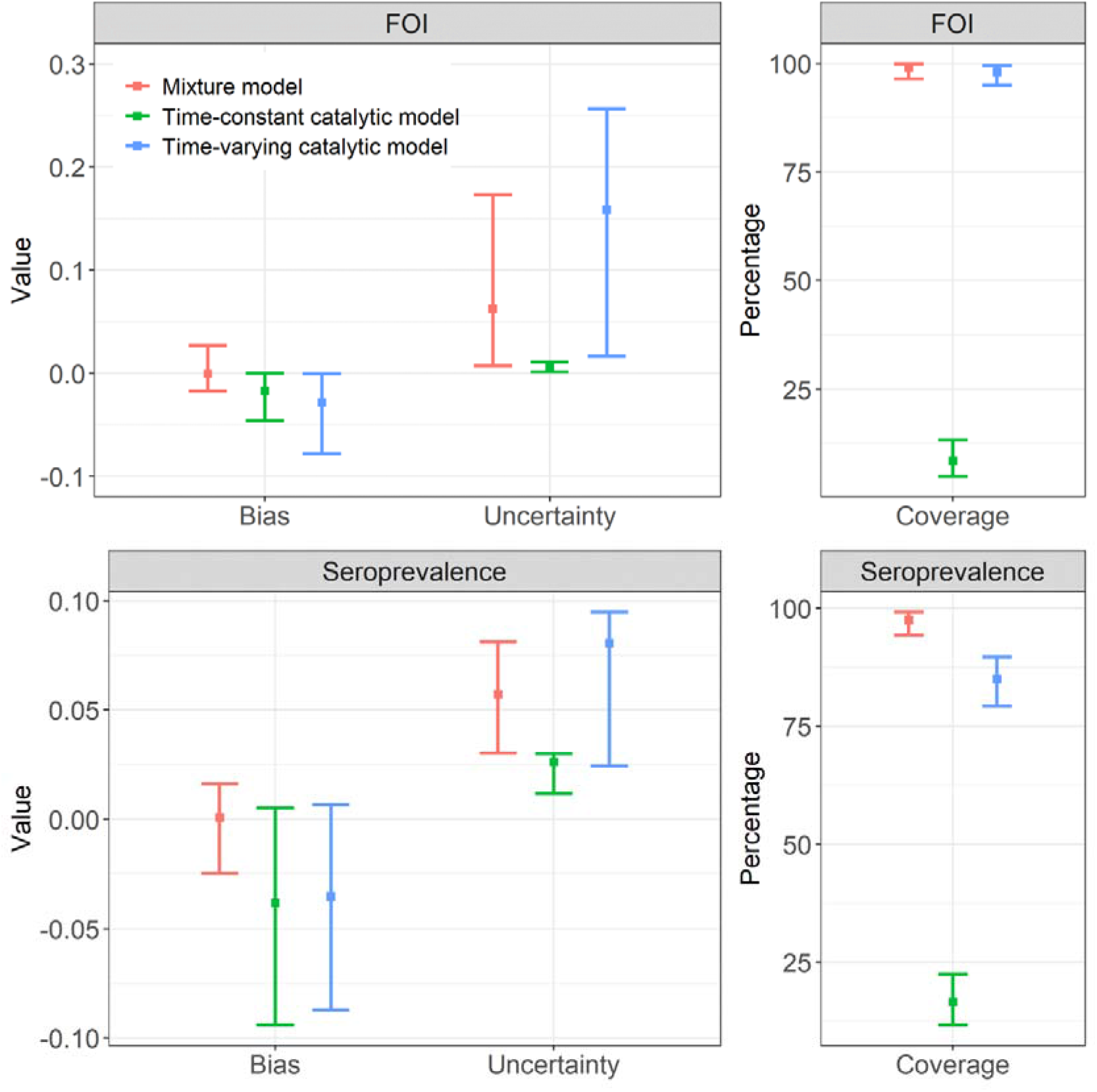
Bias, coverage, and degree of uncertainty for seroprevalence and force of infection (FOI) estimates using catalytic and mixture models fitted to simulated datasets (Dataset C). Bias is calculated as the estimated parameter value minus the true parameter value. Uncertainty is the width of the 95% Confidence intervals (CIs) around the central estimates, calculated using the bootstrap method. The coverage is the percentage of simulations where the estimated CIs contained the true values. For the bias and the uncertainty, the mean and 95% CI across the 200 simulations are given. For the coverage, the 95% exact binomial CI are given. Note that the y-axis limits differ for each panel.

The catalytic model under both assumptions underestimated the FOI and seroprevalence, particularly at higher values (Figure 1 and Figure 2). High titre misclassification error rates were positively associated with increased parameter estimate bias in the catalytic models (S7 Figure). As expected, parameter estimate bias was reduced when we fit the catalytic models to the simulated ‘true serostatus’ instead of classifying the titres using optimised thresholds: the FOI was then correctly estimated for 99.5% (199/200) and 48% (96/200) of simulations by the time-varying and time-constant FOI catalytic models, respectively.

**Figure 2:**
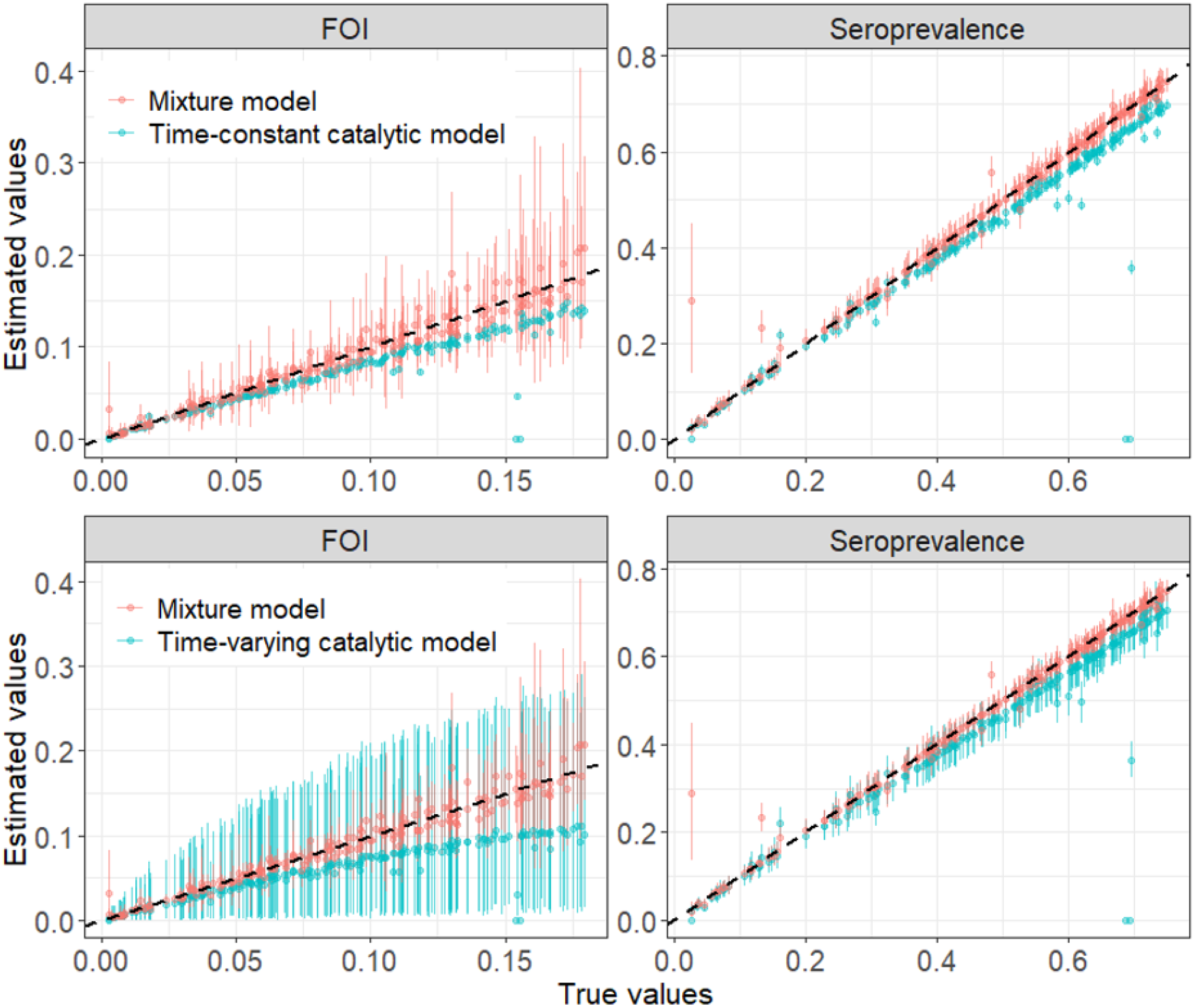
True versus estimated seroprevalence and force of infection (FOI) values from the mixture and catalytic models fitted to the simulated datasets (Dataset C). The catalytic model was run under the assumption that the FOI was time-constant or time-varying. The 95% Confidence Intervals for the estimated values were calculated using a bootstrap method and are shown here as error bars; the point denotes the central estimate.

### Long Xuyen, Vietnam data

When we applied the mixture model (Figure 3) to the data from Long Xuyen, Vietnam, the total population-level seroprevalence estimates ranged from 16.3% (95% CI 13.8%-18.8%) in 2004 to 37.6% (24.9%-40.3%) in 2005-2006. The seroprevalence estimates from the time-constant and time-varying catalytic models were consistent with each other, with the latter ranging from 18.9% (16.3%-21.7%) in 2006-2007 to 29.9% (26.2%-33.7%) in 2008-2009. The seroprevalence estimates from all three models were consistent (as determined by the 95% CIs) for 4 out of 6 datasets (Figure 4, S4 Table). The general trend in the age-specific seroprevalence estimates, specifically for Datasets A-2:A-5, differed significantly between the mixture model and the catalytic models, with the mixture model estimating higher seroprevalence at the older ages (Figure 5).

**Figure 3:**
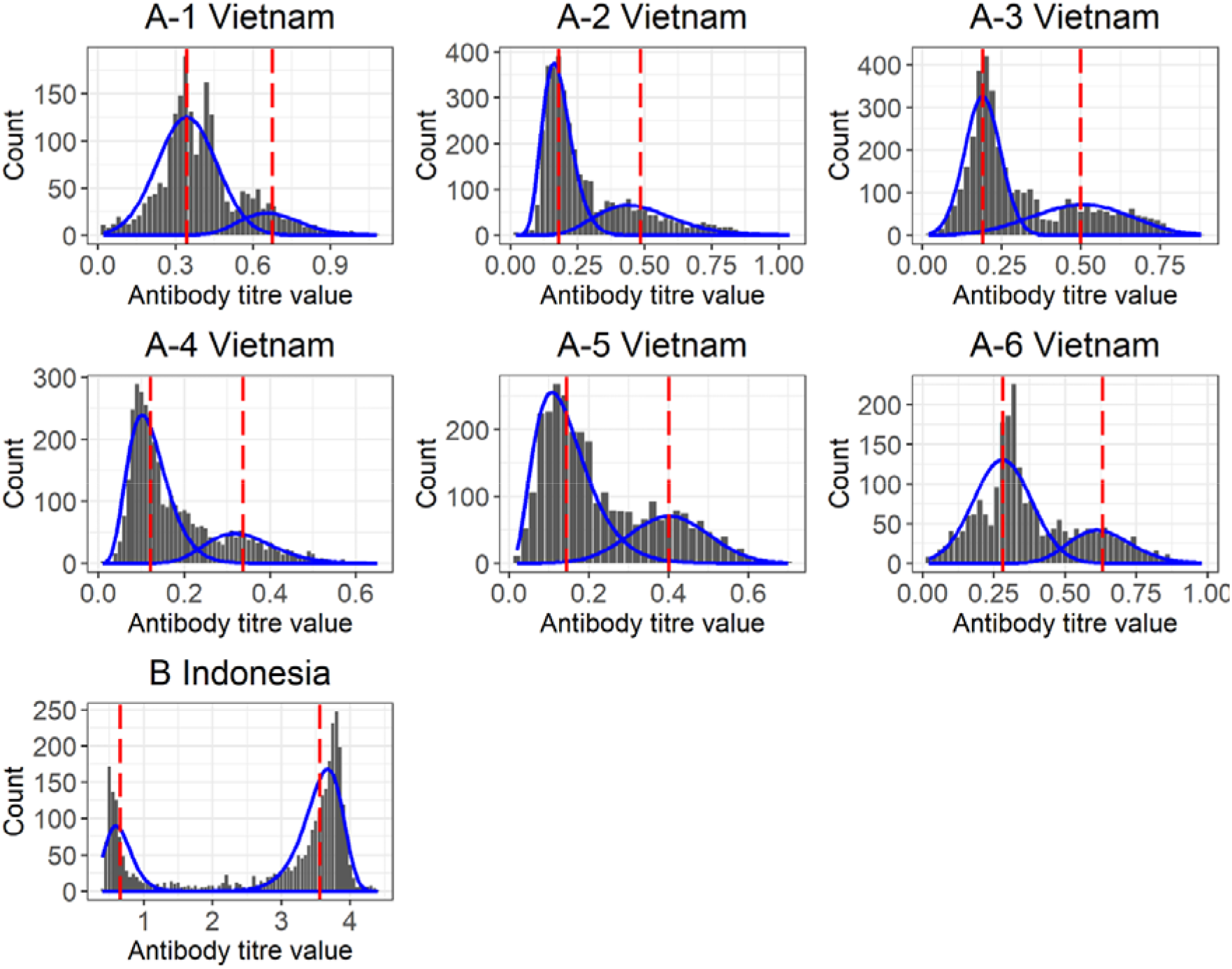
Mixture model fitted to the Vietnamese (A1:A6) and Indonesian (B) datasets. The distribution of log(titre+1) is shown in dark grey, the fitted mixture model is shown in blue, and the red dashed lines represent the mean antibody titre of each component of the fitted mixture model (*μ*_*s*_ and *μ*_*I*_ for the seronegative and seropositive components respectively). Note that the y-axis limits differ for each panel.

**Figure 4:**
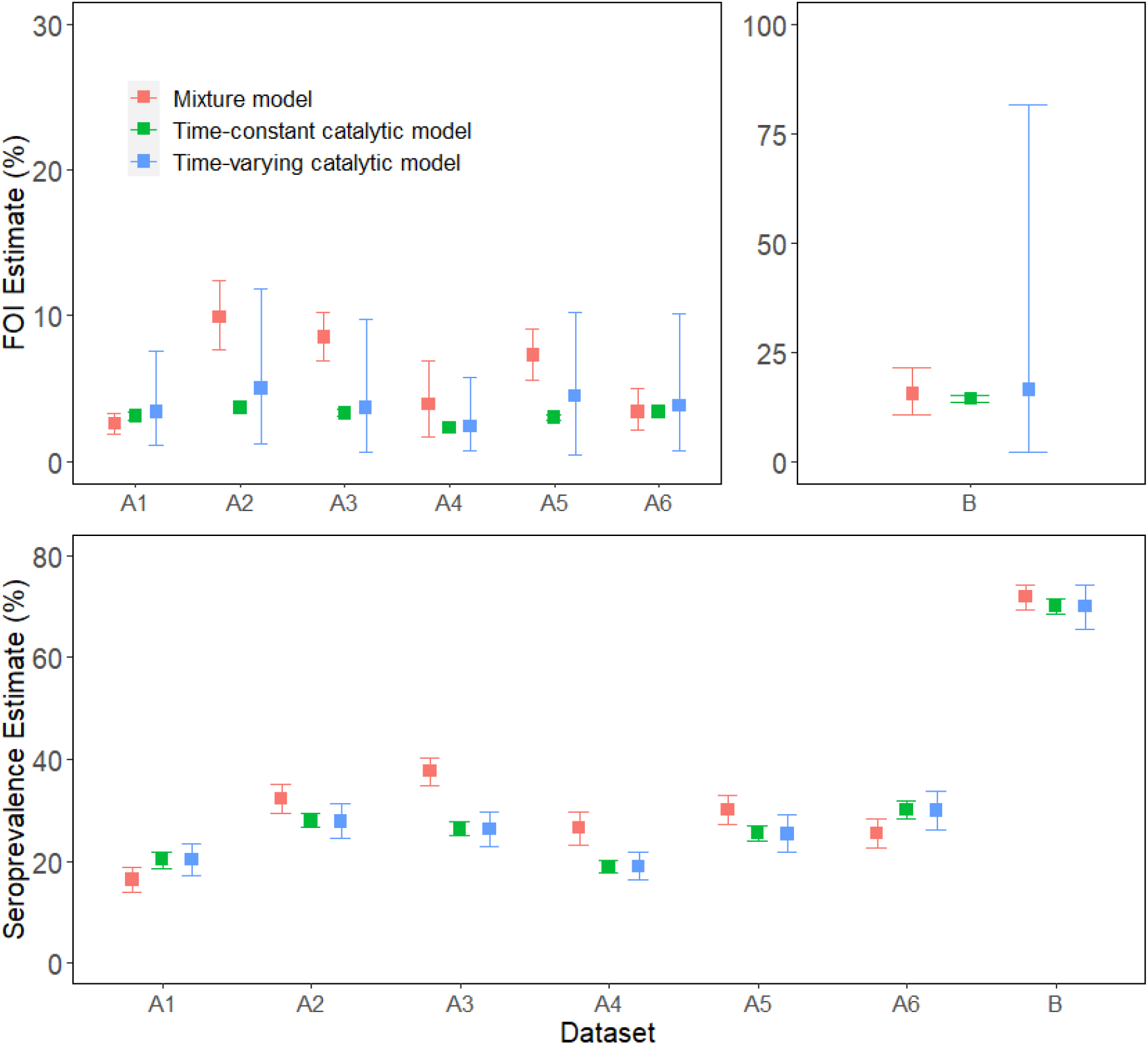
Force of infection (FOI) and total population level seroprevalence (SP) estimates from the mixture model and the catalytic models fitted to the observed data. The catalytic model was run under the time-constant and time-varying FOI assumption. The 95% Confidence Intervals (CI) which were calculated by bootstrapping for all models are given as error bars. Note that the y-axis limits differ for each panel.

**Figure 5:**
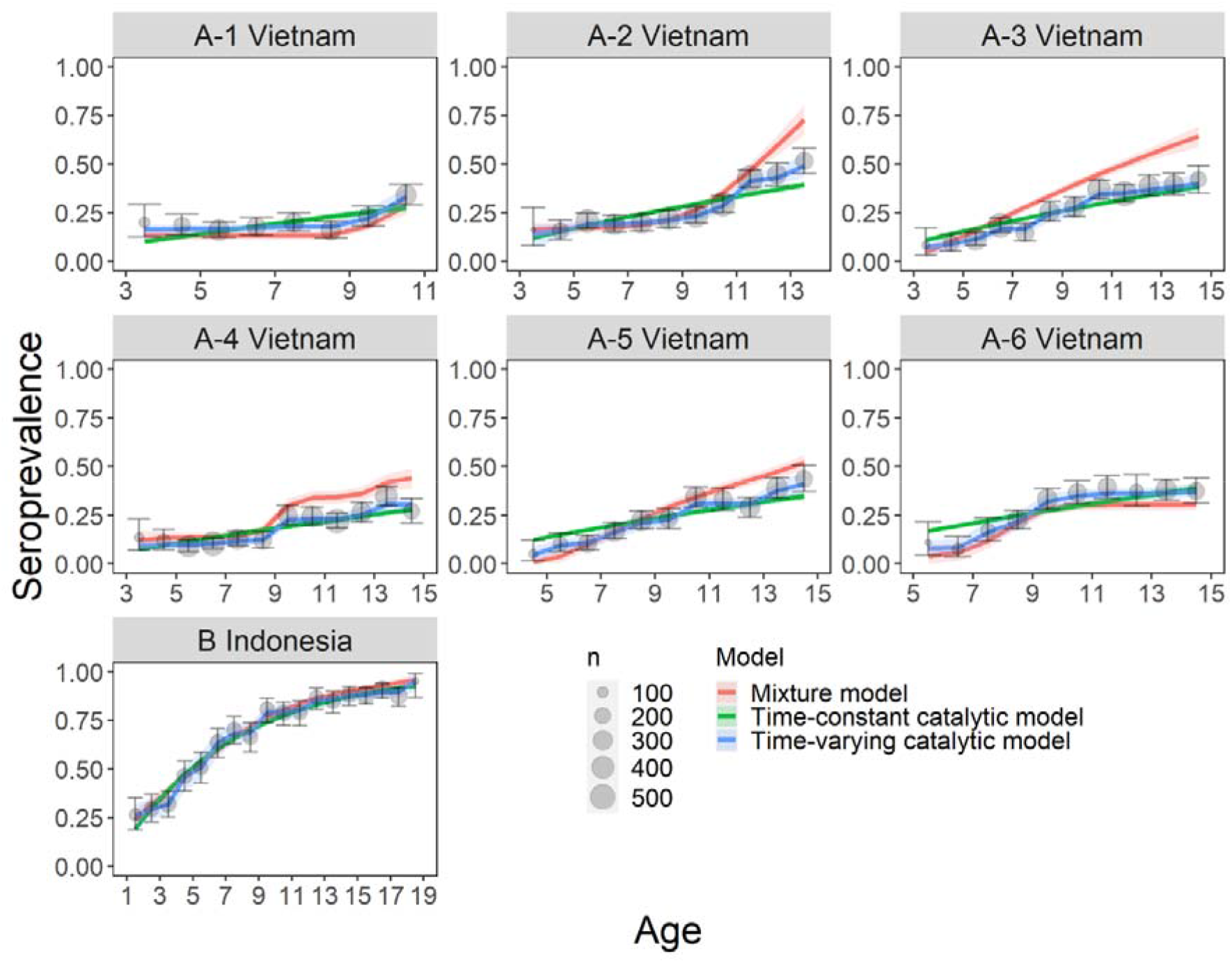
Age-specific seroprevalence estimates for the IgG data from Vietnam (Dataset A1:A6) and from Indonesia (Dataset B) aggregated across all subdistricts. Mixture model estimates are in blue, catalytic model estimates are in orange and green when applied under the assumption that the FOI is time-constant or time-varying respectively. Shading represents the 95% Confidence Intervals (CI). The grey points show the observed seroprevalence per age group calculated from the binarily classified IgG data (seropositive individuals / tested individuals), with error bars indicating the 95% exact binomial CIs. The seroprevalence data and model estimates are overlayed for the purpose of comparison. However, it is important to note that the mixture model was not fitted to the data (grey points), as the former does not depend on the titre classification. The size of the grey data points represents the number of individuals tested in each age group.

The FOI estimates ranged from 2.6% (95% CI 1.9-3.3%) in 2004 to 9.9% (7.7%-12.4%) in 2004-2005 for the mixture model and 2.4% (0.7%-5.8%) in 2006-2007 to 5.0% (1.2%-11.8%) in 2004-2005 for the time-varying catalytic model. The FOI estimates from the mixture model versus the time-varying catalytic model were consistent for 6 out of 6 datasets, and versus the time-constant catalytic model they were consistent for 3 out of 6 datasets (Figure 4, S4 Table). There is a higher degree of uncertainty around the estimates from the catalytic model when assuming a time-varying FOI compared to the time-constant FOI assumption (Figure 4). We observe greater differences in the estimates from each model when comparing the age-specific FOI as opposed to the averaged total FOI (S9 Figure).

### Indonesian data

The mixture and catalytic models fitted to the Indonesian data produced consistent FOI, total population seroprevalence and age-specific seroprevalence estimates. The FOI was estimated at 15.4% (95% CI: 10.6%-21.3%), 14.3% (13.6%-15.0%) and 16.4% (2.2%-81.4%), and the seroprevalence at 71.8% (69.4%-74.1%), 70% (68.6%-71.4%) and 70.0% (65.5%-74.3%) by the mixture model and the time-constant and time-varying catalytic models, respectively (Figure 4, S4 Table).

## Discussion

In our simulation study, the mixture model produced less biased estimates of FOI and seroprevalence than the catalytic models (Figure 1). The catalytic models consistently underestimated the parameters, particularly at higher values (Figure 2). Increased bias in the catalytic model estimates was associated with increased serostatus misclassification (S7 Figure). Serostatus misclassification occurred more often in simulations where the mean log(titre + 1) for the susceptible/seronegative component was higher and/or the mean log(titre + 1) for the infected/seropositive component was lower (S6 Figure), indicating greater overlap between the distributions of the two components. It is worth noting that the apparent poor performance of the catalytic model is directly related to our choice of simulated parameter values, and we would expect better performance when using parameters that would have given clearer bimodal titre distributions, where serostatus misclassification would have been lower.

Differences in the degree of overlap between components in real serological datasets (S8 Figure) reflect differences in transmission intensity and variable degrees of spatiotemporal heterogeneities in the risk of infection (3,37). In datasets collected from areas which experience hyperendemic DENV circulation, one expects greater separation between the titre components because most seropositive individuals likely have had multiple infections, translating to higher antibody titres. In these instances, catalytic and mixture models are expected to produce more similar estimates of FOI and seroprevalence as fewer samples are misclassified during the qualitative, binary classification of the data needed to calibrate catalytic models (22,38). This is consistent with the reduced variability we observe in our FOI and seroprevalence estimates from each model when they were applied to serological data from Indonesia compared to Vietnam, where the former had higher separation of titre distributions (Figure 3, Figure 4, S4 Table).

The FOI for Indonesia was estimated at 15.4% (95% CI: 10.6%-21.3%), 14.3% (13.6%-15.0%) and 16.4% (2.2%-81.4%), and the seroprevalence at 71.8% (69.4%-74.1%), 70% (68.6%-71.4%) and 70.0% (65.5%-74.3%) by the mixture model and catalytic model under the time-constant and time-varying FOI assumptions, respectively. The FOI estimates are consistent with previously published results from catalytic models fitted to case-notification data from 2008-2017 in Jakarta, Indonesia (13.0%, 95% CI: 12.9-13.1%) (12). The seroprevalence estimates are consistent with previously published results using time-constant catalytic models applied to the same serology dataset (Dataset B) (31,39). Our results show that the mixture and catalytic models do not significantly differ in their FOI and seroprevalence estimates in this setting.

The mixture model applied to the six datasets from Vietnam produced more variable estimates (FOI range = 2.6%-9.9%, seroprevalence range = 16%.3-37.6%) than the catalytic models (FOI range = 2.3%-3.7% and 2.4%-5.0%, seroprevalence range = 19.0%-30.0% and 18.9%-29.9% under the assumption of a time-constant or time-varying FOI respectively). The estimates from the mixture model tended to exceed those obtained from the catalytic models (Figure 4, S4 Table). Given the negative bias observed for the catalytic models in our simulation study, we expect the higher mixture model estimates to be more accurate for the Vietnamese setting. The higher FOI estimates are closer to previously published estimates of 11.7% (95% CI: 10.8-12.7%) (40) using data collected from Binh Thuan province in 2003, and 14.2% (95% CI: 12.9-18.4%) using data collected from Dong Thap province in 1996-7 (19). Previous work has demonstrated large spatiotemporal heterogeneity of DENV in Vietnam (37).

The variance between our mixture and catalytic model estimates for Vietnam is greatest in the age-specific seroprevalence and FOI estimates (Figure 5, S9 Figure). As expected, the time-varying catalytic model and the mixture model (which implicitly models FOI as time-varying), are better able to capture the age-specific seroprevalence than the time-constant catalytic model. However, the time-varying catalytic model produced less stable estimates (wider CIs) than the time-constant catalytic model or the mixture model (Figure 1 and Figure 2).

A major advantage of the mixture model is the comparative ease with which it can be applied to serology data to estimate transmission intensity without the need to use thresholds to process the data. Furthermore, to generate robust estimates, there are fewer data requirements for mixture models than for catalytic models: in the former, the data are pooled and age is used only to calculate the age-specific mean log(titre + 1) using a spline, meaning that there are no constraints on the number of participants per age category. However, it is important to consider the bias that will be introduced if the mixture distributions fit the titre data poorly (38). Biggs *et al*. fit mixture distributions to DENV antibody titre data in the Philippines to develop a framework distinguishing between post and primary DENV infection (29) by specifying a three-component mixture for seronegative, seropositive with a primary infection and seropositive with post-primary infections. In future work, it would be of interest to explore the FOI estimates obtained on the Vietnamese datasets with a similar three-component mixture model.

Our results suggest that mixture models represent a good alternative to catalytic models to quantify DENV time-varying FOI and seroprevalence from age-stratified serological data, with potentially less bias and less uncertainty. They may be particularly useful when estimating FOI in low transmission settings and/or when the study population is younger, where there is higher overlap between the component distributions (S8 Figure) so the risk of serostatus misclassification and bias introduction when using cut-off threshold methods is greater (S6 Figure and S7 Figure). We have provided code for both models to encourage further exploration and comparison of the two methods. Critically, further investigation of the use of mixture models depends on the availability of raw antibody titre data. For these reasons, we would encourage current and future seroprevalence studies on DENV as well as other infectious diseases, to publish anonymised individual-level antibody titre data where it is possible to do so.

## Data Availability

The serological data from Vietnam and Indonesia used in this work is proprietary and cannot be publicly shared. Qualified researchers may request access to the data provider. Further details on Sanofi's data sharing criteria and process for requesting access can be found at: https://www.clinicalstudydatarequest.com. The simulated data can be recreated using code that we have made available in the GitHub repository.

https://github.com/Tori-Cox/Mixture-catalytic-models

## Acknowledgements

We would like to acknowledge Sanofi Pasteur for provision of the data used in this research. We acknowledge the participants, their families and the principal investigators involved in the studies.

