## Supplementary Information for "Estimating dengue transmission intensity from serological data: a comparative analysis using mixture and catalytic models"

**Title**

**Author information**

Victoria Cox**^1^**, Megan O’Driscoll**^1,2^**, Natsuko Imai**^1^**, Ari Prayitno^3^, Sri Rezeki Hadinegoro^3^, Anne-Frieda Taurel**^4^**, Laurent Coudeville**^5^**, Ilaria Dorigatti**^1^**.

**1** MRC Centre for Global Infectious Disease Analysis, School of Public Health, Imperial College London, London, United Kingdom, **2** Department of Genetics, University of Cambridge, Cambridge, United Kingdom, **3** Department of Child Health, Faculty of Medicine Universitas Indonesia, Jakarta, Indonesia, **4** Sanofi Pasteur, Singapore, **5** Sanofi Pasteur, Lyon, France.

**Abstract**

**Background**. Dengue virus (DENV) infection is a global health concern of increasing magnitude. To target intervention strategies, accurate estimates of the force of infection (FOI) are necessary. Catalytic models have been widely used to estimate DENV FOI and rely on a binary classification of serostatus as seropositive or seronegative, according to pre-defined antibody thresholds. Previous work has demonstrated the use of thresholds can cause serostatus misclassification and biased estimates. In contrast, mixture models do not rely on thresholds and use the full distribution of antibody titres. To date, there has been limited application of mixture models to estimate DENV FOI.

**Methods**. We compare the application of mixture models and time-constant and time-varying catalytic models to simulated data and to serological data collected in Vietnam from 2004 to 2009 (N ≥ 2178) and Indonesia in 2014 (N = 3194).

**Results**. The simulation study showed greater estimate bias from the time-constant and time-varying catalytic models (FOI bias = 1.3% (0.05%, 4.6%) and 2.3% (0.06%, 7.8%), seroprevalence bias = 3.1% (0.25%, 9.4%) and 2.9% (0.26%, 8.7%), respectively) than from the mixture model (FOI bias = 0.41% (95% CI 0.02%, 2.7%), seroprevalence bias = 0.11% (0.01%, 3.6%)). When applied to real data from Vietnam, the mixture model frequently produced higher FOI and seroprevalence estimates than the catalytic models.

**Conclusions**. Our results suggest mixture models represent valid, potentially less biased, alternatives to catalytic models, which could be particularly useful when estimating FOI and seroprevalence in low transmission settings, where serostatus misclassification tends to be higher.

**Supplementary Data**

**Table S1: Details of the 30 Indonesian subdistricts where IgG titres were collected in 2014 (Dataset B).** Subdistrict ID names and details are given, with N being the number of participants in the dataset from each subdistrict [38].

| Subdistrict ID | N | Province | Regency | Subdistrict name |
| --- | --- | --- | --- | --- |
| 1 | 106 | NANGGROE ACEH DARUSSALAM | SUBULUSSALAM | SIMPANG KIRI |
| 2 | 107 | SUMATERA UTARA | MEDAN | MEDAN DENAI |
| 3 | 106 | SUMATERA BARAT | PADANG | PAUH |
| 4 | 107 | JAMBI | BUNGO | BUNGO DANI |
| 5 | 105 | LAMPUNG | LAMPUNG SELATAN | KALIANDA |
| 6 | 107 | BANTEN | TANGERANG | CIKUPA |
| 7 | 101 | BANTEN | TANGERANG | BENDA |
| 8 | 105 | DKI JAKARTA | JAKARTA SELATAN | PESANGGRAHAN |
| 9 | 107 | DKI JAKARTA | JAKARTA TIMUR | PULO GADUNG |
| 10 | 107 | DKI JAKARTA | JAKARTA BARAT | KALI DERES |
| 11 | 107 | JAWA BARAT | BOGOR | GUNUNG PUTRI |
| 12 | 107 | JAWA BARAT | BANDUNG | BANJARAN |
| 13 | 107 | JAWA BARAT | CIREBON | GUNUNG SARI |
| 14 | 106 | JAWA BARAT | BEKASI | CIKARANG UTARA |
| 15 | 107 | JAWA BARAT | BANDUNG | BOJONGLOA KALER |
| 16 | 107 | JAWA BARAT | BEKASI | BEKASI TIMUR |
| 17 | 107 | JAWA BARAT | TASIKMALAYA | SINGAPARNA |
| 18 | 107 | JAWA TENGAH | KLATEN | TRUCUK |
| 19 | 107 | JAWA TENGAH | JEPARA | PECANGAAN |
| 20 | 107 | JAWA TENGAH | TEGAL | DUKUHTURI |
| 21 | 107 | JAWA TENGAH | TEGAL | TEGAL BARAT |
| 22 | 107 | JAWA TIMUR | PONOROGO | PULUNG |
| 23 | 106 | JAWA TIMUR | BANYUWANGI | CLURING |
| 24 | 107 | JAWA TIMUR | MOJOKERTO | NGORO |
| 25 | 105 | JAWA TIMUR | SUMENEP | KALIANGET |
| 26 | 107 | JAWA TIMUR | SURABAYA | SAWAHAN |
| 27 | 107 | BALI | DENPASAR | DENPASAR SELATAN |
| 28 | 107 | KALIMANTAN TIMUR | SAMARINDA | SAMARINDA ULU |
| 29 | 107 | SULAWESI SELATAN | TORAJA UTARA | RANTEPAO |
| 30 | 107 | SULAWESI TENGGARA | KENDARI | KENDARI |

**Table S2: Parameter values used for generating 200 simulated datasets.**

| Parameter | Symbol | Uniform distribution limits |
| --- | --- | --- |
| Force of infection (FOI) | $\lambda$ | 0.001 – 0.18 |
| Mean seronegative titre | $\mu_{S}$ | 0.05 – 1.2 |
| Mean seropositive titre | $\mu_{I}$ | 2.2 – 4.0 |
| Standard deviation of the seronegative titre distribution | $\sigma_{S}$ | 0.1 – 0.8 |
| Standard deviation of the seropositive titre distribution | $\sigma_{I}$ | 0.1 – 0.8 |

**Supplementary Results**

**Table S3: Percentage of simulations where the mixture model correctly specified the seronegative and/or seropositive component of the simulated antibody titre datasets (Dataset C).** Here, n represents the number of simulated datasets out of 200.

| True seronegative distribution | True seropositive distribution | n | Correct seronegative distribution only | Correct seropositive distribution only | Correct both distributions | Incorrect both distributions |
| --- | --- | --- | --- | --- | --- | --- |
| Weibull | **Weibull** | **28** | 0% | 89% | 0% | 11% |
| Weibull | **Normal** | **27** | 0% | 67% | 0% | 33% |
| Weibull | **Gamma** | **32** | 0% | 91% | 0% | 9% |
| Normal | **Weibull** | **29** | 7% | 0% | 93% | 0% |
| Normal | **Normal** | **12** | 17% | 0% | 83% | 0% |
| Normal | **Gamma** | **17** | 0% | 0% | 100% | 0% |
| Gamma | **Weibull** | **17** | 0% | 0% | 100% | 0% |
| Gamma | **Normal** | **22** | 27% | 0% | 73% | 0% |
| Gamma | **Gamma** | **16** | 19% | 0% | 81% | 0% |

**Table S4: Force of infection (FOI) and total population level seroprevalence (SP) estimates from the mixture model and the catalytic models fitted to the observed data.** The observed data is serology data collected in Vietnam (Datasets A-1:A-6) and Indonesia (Dataset B). 95% Confidence Intervals (CI) were calculated by the bootstrap method.

| Dataset | Parameter | Mixture model  (95% CI) | Time-varying catalytic model (95% CI) | Time-constant catalytic model (95% CI) |
| --- | --- | --- | --- | --- |
| A-1 | FOI | 2.6% (1.9%-3.3%) | 3.4% (1.1%-7.6%) | 3.1% (2.8%-3.4%) |
|  | SP | 16.3% (13.8%-18.8%) | 20.3% (17.3%-23.4%) | 20.3% (18.6%-21.9%) |
| A-2 | FOI | 9.9% (7.7%-12.4%) | 5.0% (1.2%-11.8%) | 3.7% (3.5%-3.9%) |
|  | SP | 32.2% (29.3%-35.2%) | 27.8% (24.4%-31.2%) | 28.0% (26.6%-29.4%) |
| A-3 | FOI | 8.5% (6.9%-10.2%) | 3.7% (0.6%-9.8%) | 3.3% (3.1%-3.6%) |
|  | SP | 37.6% (34.9%-40.3%) | 26.2% (22.9%-29.7%) | 26.4% (25.0%-27.8%) |
| A-4 | FOI | 3.9% (1.7%-6.9%) | 2.4% (0.7%-5.8%) | 2.3% (2.1%-2.4%) |
|  | SP | 26.5% (23.2%-29.6%) | 18.9% (16.3%-21.7%) | 19.0% (17.8%-20.2%) |
| A-5 | FOI | 7.3% (5.6%-9.1%) | 4.5% (0.5%-10.2%) | 3.0% (2.8%-3.2%) |
|  | SP | 30.1% (27.3%-33.0%) | 25.3% (21.7%-29.1%) | 25.5% (24.0%-27.0%) |
| A-6 | FOI | 3.4% (2.2%-5.0%) | 3.8% (0.7%-10.1%) | 3.4% (3.1%-3.6%) |
|  | SP | 25.4% (22.5%-28.3%) | 29.9% (26.2%-33.7%) | 30.0% (28.2%-31.8%) |
| B | FOI | 15.4% (10.6%-21.3%) | 16.4% (2.2%-81.4%) | 14.3% (13.6%-15.0%) |
|  | SP | 71.8% (69.4%-74.1%) | 70.0% (65.5%-74.3%) | 70.0% (68.6%-71.4%) |

**Figure S5: True versus estimated parameter values from the mixture model fitted to the simulated datasets (Dataset C).** The estimated parameters are the mean log(titre + 1) value of the seronegative/susceptible (S) and seropositive/infected (I) components (muS ($\mu_{s}$) and muI ($\mu_{I}$) respectively) and the corresponding standard deviations (sdS ($\sigma_{s}$) and sdI ($\sigma_{I}$)). Red indicates the estimates where the true parameter value was not captured by the estimates (i.e., the 95% Confidence Interval of the estimate did not contain the true value). Note that the y-axis limits differ for each panel.

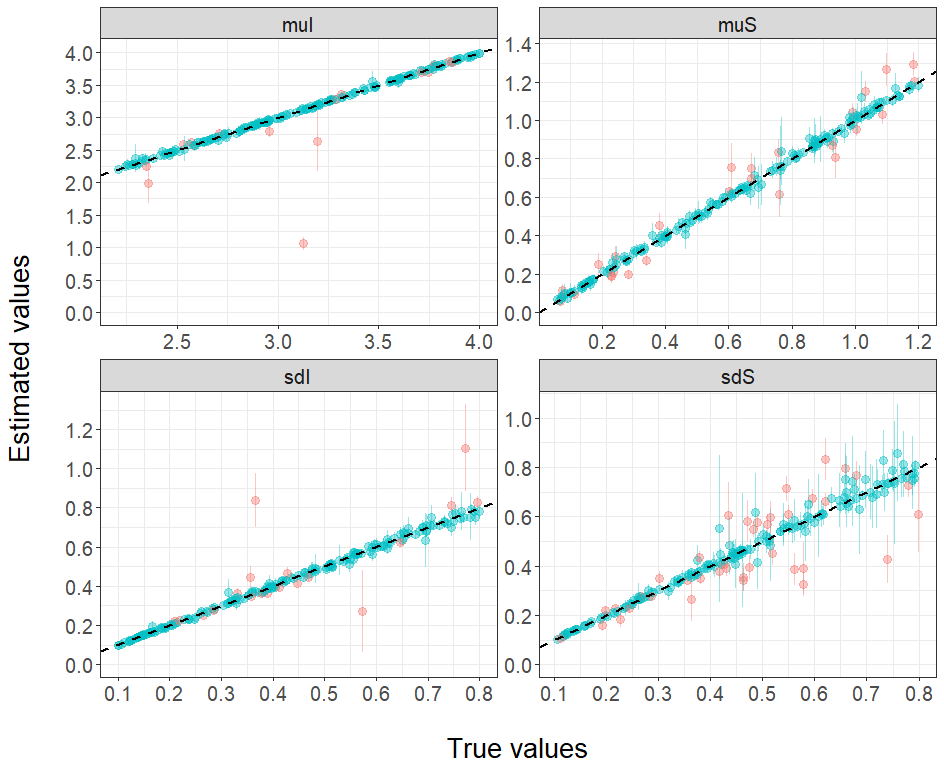

**Figure S6: Association between the true component mean titre values in Dataset C versus the serostatus misclassification error.** The x-axis shows the difference between the true mean log(titre + 1) value of the seronegative ($\mu_{s}$) and the seropositive component ($\mu_{I}$) for each realisation over 200 simulated datasets. The titres are classified as seropositive or seronegative using realisation-specific optimised titre thresholds. The linear regression line and corresponding 95% Confidence Intervals are shown, as well as the correlation coefficients (*R*) and p-value for the slope coefficient (*p*).

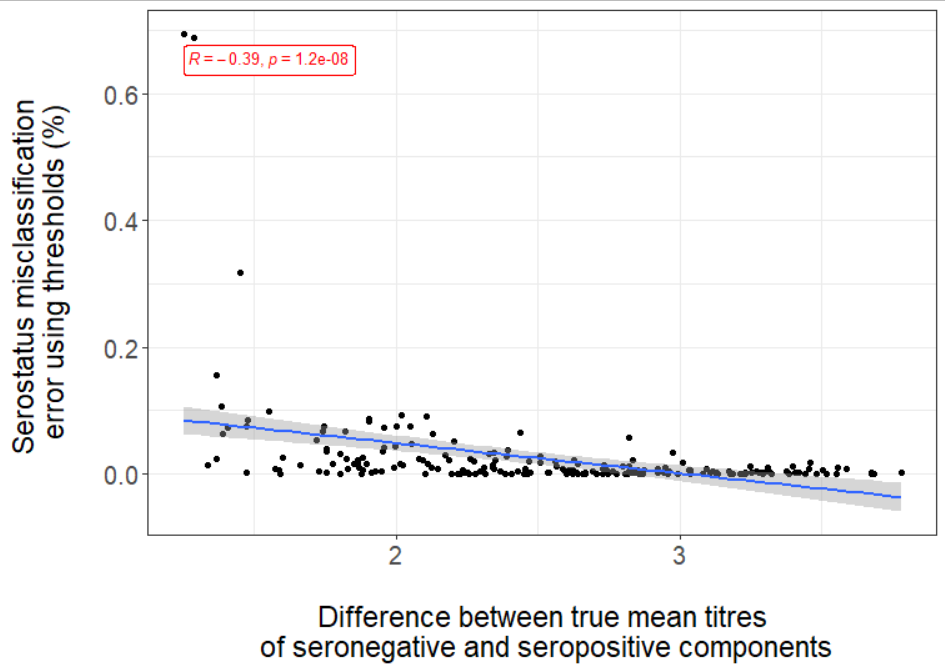

**Figure S7: Serostatus misclassification versus catalytic model estimate bias.** The absolute bias of the realisations over 190 simulated datasets (10 outliers with misclassification error > 10% were removed) are plotted against the mean serostatus misclassification error. The serostatuses of the titres are classified as seropositive or seronegative using realisation-specific optimised titre thresholds. Absolute bias is calculated as the absolute value of the estimated value – true value for the force of infection (FOI) and seroprevalence. The linear regression lines and corresponding 95% Confidence Intervals are shown, as well as the correlation coefficients (*R*) and p-value for the slope coefficient (*p*).

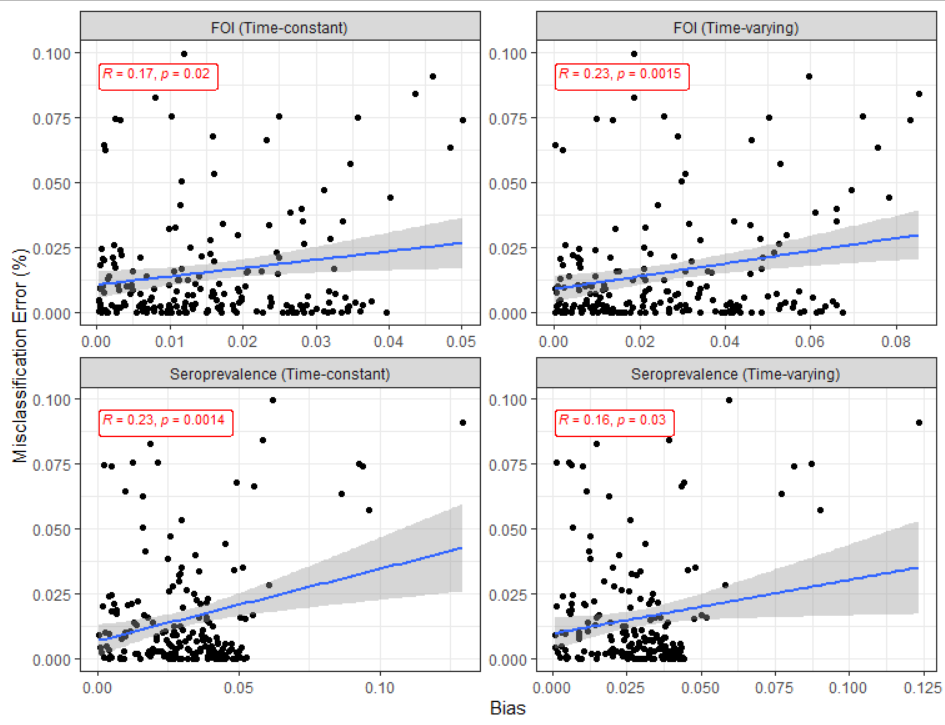

**Figure S8: Association between the estimated force of infection (FOI) and component distribution parameters in the real serology datasets (Datasets A-1:A-6 and B).** FOI estimates from the mixture model (y-axis) plotted as a function of the difference between the mean log(titre + 1) for the susceptible/seronegative component (muS ($\mu_{s}$)) and the infected/seropositive component (muI ($\mu_{I}$)) (x-axis).

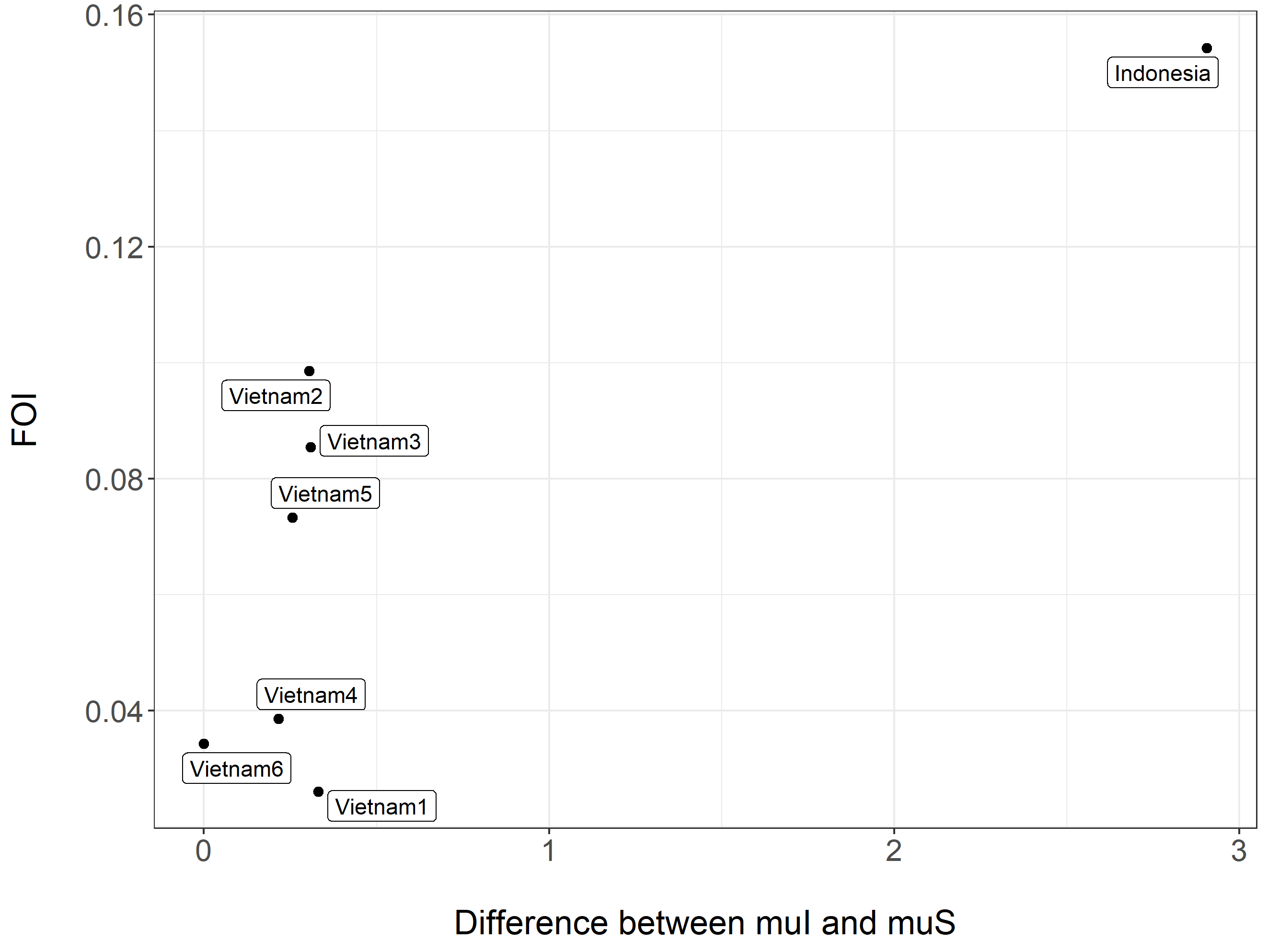

**Figure S9: Force of infection (FOI) estimates across time.** Age-specific FOI estimates from the catalytic and mixture models. 95% Confidence Intervals were calculated by bootstrapping. The estimates for each age class, *a*, represent either age-specific FOI (or equivalently the time-specific FOI *a* to *a+1* years ago). The data was collected in 2004, 2004-2005, 2005-2006, 2006-2007, 2007-2008, 200-2009 and 2014 for Datasets A-1:A-6 and B, respectively. Therefore, the FOI at age-class 3 years estimated using Dataset A-1, could be interpreted as the time-specific FOI experienced by individuals in 2001.

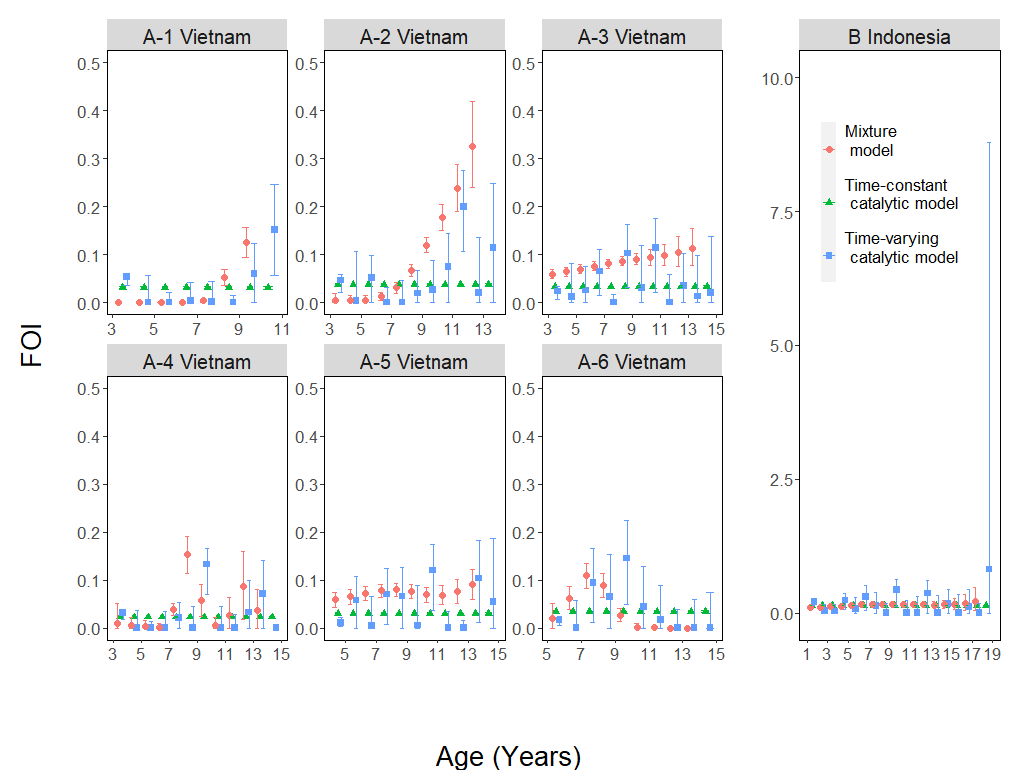
